# Prediction of Pediatric No-Show Rates Before and During the COVID-19 Pandemic Utilizing Machine Learning

**DOI:** 10.1101/2024.11.23.24317836

**Authors:** Quincy A. Hathaway, Naveena Yanamala, TaraChandra Narumanchi, Janani Narumanchi

## Abstract

**Background:** Patient no-shows significantly disrupt pediatric healthcare delivery, highlighting the necessity for precise predictive models, especially during the dynamic shifts caused by the SARS-CoV-2 pandemic. In outpatient settings, these no-shows result in medical resource underutilization, increased healthcare costs, reduced access to care, and lower clinic efficiency and productivity.

**Methods:** The objective is to develop a predictive model for patient no-shows using data-driven techniques. We analyzed five years of historical data retrieved from both a scheduling system and electronic health records from a general pediatrics clinic within the WVU Health systems. This dataset comprises a total of 209,408 visits from 2015 to 2018, 82,925 visits in 2019, and 58,820 visits in 2020, spanning both pre-pandemic and pandemic periods. The data includes variables such as patient demographics, appointment details, timing, hospital characteristics, appointment types, and environmental factors.

**Results:** Our XGBoost model demonstrated robust predictive capabilities, notably outperforming traditional "no-show rate" metrics. Precision and recall metrics for all features were 0.82 and 0.88, respectively. Receiver Operator Characteristic (ROC) analysis yielded AUCs of 0.90 for all features and 0.88 for the top 5 predictors when evaluated on the 2019 cohort. Furthermore, model generalization across racial/ethnic groups was also observed. Evaluation on 2020 telehealth data reaffirmed model efficacy (AUC: 0.90), with consistent top predictive features.

**Conclusions:** Our study presents a sophisticated predictive model for pediatric no-show rates, offering insights into nuanced factors influencing attendance behavior. The model’s adaptability to evolving healthcare landscapes, including telehealth, underscores its potential for enhancing clinical practice and resource allocation.

## Introduction

Patient no-shows, defined as a patient failing to attend a scheduled appointment without prior notification, vary widely across healthcare settings, ranging from 12% to 80% (Marbouh et al., 2020). No-shows are a major logistical and economic challenge for clinics and hospital systems, leading to significant revenue losses. No-shows cost the U.S. healthcare system more than $150 billion a year and individual physicians an average of $200 per unused time slot (Gier, 2017). Whether patients show up or not, healthcare organizations and medical practices must still pay their staff and cover expenses of resources/facilities. For the provider, no-showed appointments decrease the volume of medical care that can be given. For patients, the increasing lengths of time needed to schedule follow-up appointments can be prohibitive of receiving proper care (Ansell et al., 2017). Patients who failed to keep an appointment were up to 70% more likely not to return within 18 months, with older patients experiencing more chronic illnesses and more likely not to return to their physicians’ offices after missing just one appointment (Hayhurst, 2019).

Several studies have modeled strategies for reducing patient wait times for clinic appointments through artificial intelligence (AI)-enabled technologies and solutions (Salazar et al., 2022). These machine and deep learning approaches have utilized random forest (Qureshi et al., 2021;Salazar et al., 2021), logistic regression (Moharram et al., 2021;Qureshi et al., 2021), gradient boosting (Daghistani et al., 2020;Fan et al., 2021), ensemble-based models (Ahmadi et al., 2019;Alshammari et al., 2021), deep neural networks (Alshammari et al., 2020;Srinivas and Salah, 2021), and various other approaches to predict patient no-shows. Studies have also examined the type of visit, whether primary care or specialty clinic, and varying demographic populations (e.g., United States, Saudi Arabia, Brazil, etc.) (Salazar et al., 2022). While outpatient scheduling with a healthcare provider is traditionally thought to be the largest percentage of no-showed appointments, the use of no-show algorithms could also predict patient readmissions (Shameer et al., 2017) and attrition from diagnostic visits (i.e., imaging and laboratory studies) (Chong et al., 2020). These individualized approaches boost patient/provider satisfaction (Mohammadi et al., 2018) and allow for a more efficient model of healthcare.

The factors affecting patient no-shows can vary slightly between studied populations but can be generalized into three major categories: 1) modifiable and unmodifiable patient characteristics (e.g., age, sex, race/ethnicity, BMI), 2) type of appointment scheduled (i.e. primary care or specialty clinic), and 3) patient behavior (e.g., previous no-show rate, time-to-appointment, travel distance, weather, etc.) (Salazar et al., 2022). Previous research indicates patients who miss appointments tend to be of lower socioeconomic status, often have a history of failed/no-show appointments, government-provided health benefits, and psychosocial problems who are less likely to understand the purpose of the appointment (Ellis et al., 2017;Daghistani et al., 2020;Marbouh et al., 2020). In addition to forgetting appointments, issues such as trouble getting off work, finding childcare, transportation, and cost can also limit patient compliance for an appointment. No-show rates also increase with increasing time between scheduling and the actual appointment (Festinger et al., 2002). In pediatrics, few studies have examined how no-show rates can be predicted using machine learning (Chen et al., 2020;Liu et al., 2022), with no pediatric studies exploring how the SARS-CoV-2 pandemic affects the ability of no-show rates to handle virtual appointments.

Our study, assess the ability of a machine learning model to predict no-show rates in a pediatric population both before and during the SARS-CoV-2 pandemic. We utilized electronic medical record (EMR) data for patients, including features related to modifiable and unmodifiable patient characteristics, appointment type, and patient behavior. We build our model on pre-pandemic outpatient appointment data and utilized pre-pandemic and pandemic derived external validation sets. We were able to effectively predict pediatric no-show rates in our validation sets and further explored the role of race/ethnicity in no-show rate prediction.

## Methods

### Ethical Approval

The study was approved by West Virginia University Institutional Review Board.

### Study Population & Data Preprocessing

Data from medical appointments were gathered from an outpatient clinic within a prominent academic pediatric hospital (West Virginia University Hospitals), aimed at enhancing quality improvement efforts. This dataset encompasses appointments across all clinic departments on main campus and satellite, outreach locations and visit types, including information on healthcare providers and weather/environmental conditions. To facilitate analysis, categorical data were transformed into multiple binary variables. For instance, the original "day of the week of the appointment" feature, ranging from 1 to 7 (representing Sunday to Saturday), was converted into seven binary indicators, each indicating the appointment’s occurrence on a specific day. Numerical features were normalized to a range of 0 to 1. The labels indicating appointment outcomes were binary, with 1 indicating a no-show and 0 denoting attendance. Notably, a single patient may have multiple records due to multiple appointments. The patient demographic largely comprises children, often accompanied by their parents or caregivers to appointments.

### Machine Learning Algorithm Development

Our current model leverages pediatric appointments from 2015-2018 (training/testing), 2019 (validation), and 2020 (external holdout) that were scheduled at West Virginia University Outpatient Clinics under the West Virginia University Hospital Systems in West Virginia. The datasets consist of 209,408 (2015-2018), 82,925 (2019), and 58,820 (2020) patient appointments. A total of 46 features were collected that included demographic factors, time of appointment, hospital variables, type of appointment scheduled, and environmental conditions.

Our machine learning model relies on a XGBoost framework that allows for adaptable weighting of variables and hyperparameter optimization [30]. To predict patient no-shows, we implemented the XGBoost algorithm using the XGBClassifier, chosen for its robust performance on complex datasets. The classifier was configured with a gradient boosting ‘gbtree’ booster and a base score of 0.5. We set the learning rate to 0.3 and the maximum depth of trees to 6, ensuring the model was sufficiently detailed yet avoided overfitting. Each tree node considered all features due to a subsample rate and colsample parameters set to 1. The model utilized 100 estimators, with the optimization objective set to binary logistic, tailored for binary classification tasks.

For regularization, which helps reduce model complexity and enhance performance, we used a lambda value of 1 and an alpha value of 0. Handling missing values automatically, the model treated missing data points as NaNs, allowing for flexibility in dealing with incomplete records. The model operated under a binary logistic objective, focusing on the probability of no-show events. A total of four parallel jobs were run (***n_jobs*** set to 4), exploiting multi-core processing to expedite computation. The random state was anchored at 0 to ensure consistency and reproducibility across model runs. We employed an automatic predictor setting, which optimally selected the most efficient prediction method based on the data structure. The tuning of hyperparameters like ***max_depth, min_child_weight, subsample, and colsample_bytree*** was conducted through a methodical grid search to identify the optimal balance, enhancing model effectiveness without overfitting. This methodological approach was geared towards developing a robust predictive model that could effectively forecast patient no-show probabilities in pediatric outpatient settings, considering various patient and environmental factors.

### Performance Evaluation Metrics

The model’s performance was evaluated using standard metrics: accuracy, precision, recall, and area under the ROC curve (AUROC). The ROC curve plots the true positive rate (sensitivity) against the false positive rate (1-specificity) at different thresholds ranging from 0 to 1. The prediction scores (i.e., the predicted probabilities of no-shows) are compared at each threshold. A higher AUROC value (closer to 1) indicates better prediction quality. Similarly, the precision-recall curve (PRC) and area under the PRC (AUPRC) were calculated. Precision measures the accuracy of positive predictions, while recall measures the model’s ability to identify all actual positives. These metrics collectively provide insights into the model’s predictive accuracy and effectiveness.

### SHAP Feature Analysis

We employed SHAP (SHapley Additive exPlanations) feature analysis to interpret the predictive model’s behavior and understand the importance of each feature in making predictions. SHAP values provide insights into how individual features contribute to the model’s output. By decomposing the model’s output for each prediction, SHAP enables us to understand the impact of each feature on the prediction outcome. Particularly, SHAP summary plots was generated to visualize the overall feature importance and understand the relationship between specific features and the predicted outcome. Positive SHAP values indicate a feature that contributes to increasing the prediction, while negative values suggest a feature that decreases the prediction.

### Intellectual Property/Data Availability

Our machine learning algorithm is covered by a provisional patient filed between Aspirations LLC and West Virginia University. This is distinctly unique from other filed patients, including US0150242819A1 [2015] – utilizing advanced statistical techniques with no indication of accuracy or performance of the models. US20110208674A1[ 2010] – a similar concept but within a ticket booking system. WO2018058189A1 [2016] – describes a supervised learning module that targets overbooking strategies, rather than uniquely identifying patients who are at risk of no-showing their appointment. Due to confidentiality agreements and the proprietary nature of the technology, the underlying data supporting this work is not publicly available.

### Statistics

Baseline characteristics of the dataset were analyzed to provide insights into the demographic, clinic-based, insurance and appointment-related attributes of the patient population. Descriptive statistics, including measures of central tendency and dispersion, were computed for numerical variables such as age and appointment duration etc. Categorical variables, such as gender and appointment type, were summarized using frequency distributions.

To identify factors influencing the likelihood of patient no-shows, univariate and multivariate analyses were conducted. Univariate analyses involved assessing the association between each individual predictor variable and the outcome variable (completed vs. show) using appropriate statistical tests such as chi-square tests for categorical variables and t-tests or ANOVA for continuous variables.

## Results

### Baseline Characteristics and Influence of Variables on No-show Rates

We retrospectively collected records from 161,822 hospital appointments made by 19,450 patients between January 1^st^, 2015 and December 31^st^, 2019 at pediatric clinics of all specialties in West Virginia University Hospitals [WVUH]. From our experience the main factors driving the no-show rate were the days until the scheduled appointments. The longer the interval, the less likelihood of the appointment being completed. We also noted that same day appointments had low likelihood of patient no-shows. If the appointment was canceled or rescheduled that was another important predictive factor resulting in the no-shows. Interestingly full-time employment status of the parent had a positive impact on the adherence to the appointment in our pediatric clinics. Thos with previous history of no-shows tend to have more chances of missing future appointments. Though it was not a major factor? but we noted that some pediatric specialties had higher chances of completing appointments for example Cardiology, Nephrology while others had higher chances of experiencing patient no-shows for example Hematology, Neurology, Gastroenterology.

Gender, weather, race or ethnicity, language preference were not significantly impacting the status of a no-show for a scheduled appointment.

### Performance of model predictions of no-shows

The machine learning model developed the no-show prediction probabilities. Table 2 highlights the superior prediction capacity of patient no-shows when using all 46 features collected (Precision: 0.82, Recall: 0.88) as well as the top 5 predictive features (Precision: 0.81, Recall: 0.84) in the validation dataset. This is further captured by the Receiver Operator Characteristic (ROC) Area Under the Curve (AUC) for all features (AUC: 0.90) and top 5 features (AUC: 0.88) (Figure 1). Additionally, we used the basic “no-show rate” alone to compute the likelihood of a patient being compliant with their visit (AUC: 0.64) (Figure 1). This “no-show rate” is a simple frequency: (total visits the patient has no-showed) / (total visits the patient has attended + total visits the patient has no-showed). This frequency is commonly employed by EMR systems to provide a baseline estimation if double booking or other alternative scheduling procedures should be enacted. To test if our algorithm can provide unbiased predictions across racial/ethnic groups, we subset the data. While Caucasians make up the primary patient population, our algorithm can efficiently generalize to other racial and ethnic populations, even when underrepresented (Table 3). From our analyses in the pediatric population, the no-show rate alone was insufficient to effectively predict patient compliance with their appointment. Additionally, features that were most important to the construction of the model were not within a single category, highlighting the complexity in interpretating of patient no-shows (Figure 2). The top five most predictive features extended across demographic factors, time of the appointment, hospital variables, and type of appointment.

**Figure 1.**
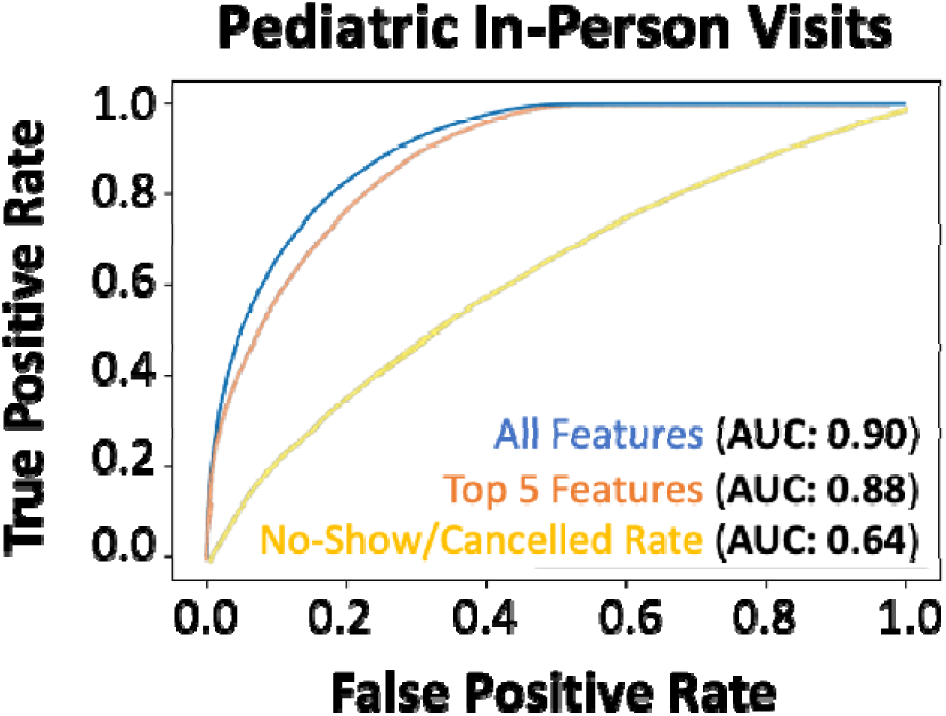
Performance of the machine learning (XGBoost) model for predicting no-shows in the hold-out test set. AUROC of xgboost machine learning model all features (blue line), only the top-5 features (orange line) and the direct no-show/cancel rate (yellow line) in predicting no-shows for 2019 hold-out validation dataset.

**Figure 2.**
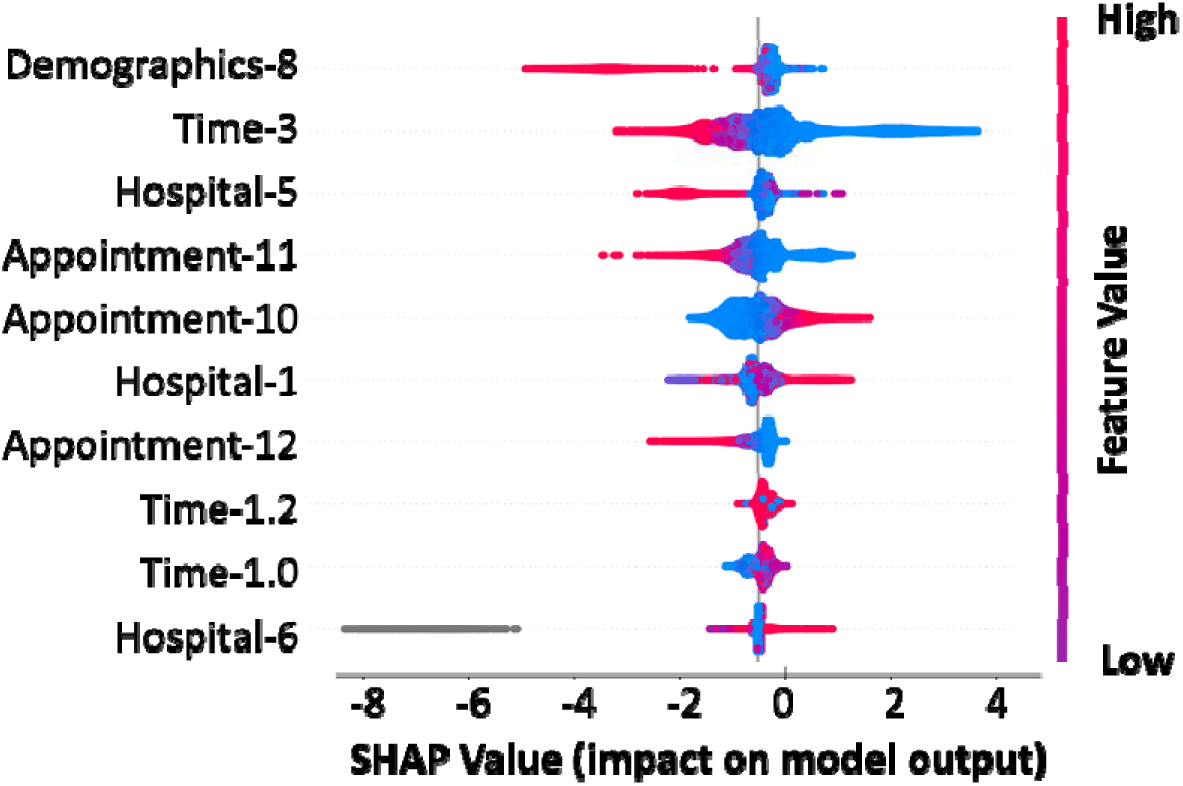
Feature importance in predicting no-shows. SHAP summary plot for the top 10 features contributing to the XGBoost model. Each line represents a feature, and the abscissa is the SHAP value. Red dots represent higher feature values, blue dots represent lower feature values and grey dots represent missing values.

**Table 1:** Details of the Study Population and influence of the variables across the show and no-show appointments.

| Parameter | Completed Appointment<br>[95% CI] (n=116,442) | Not Completed Appointment<br>[95% CI] (n=93,202) | P-Value | Cohen (Effect Size) |
| --- | --- | --- | --- | --- |
| **Patient's Age (years)** | 11.56 [11.49-11.62] | 11.27 [11.2-11.33] | <0.001 | 0.0267 |
| **Gender (% Male)** | 61478 (52.80%) | 48409 (51.94%) | <0.001 | 0.0172 |
| **Race Ethnicity (% Caucasian)** | 108668 (93.32%) | 86103 (92.38%) | <0.001 | 0.0377 |
| **English Fluency (% Fluent)** | 114644 (98.46%) | 91307 (97.97%) | <0.001 | 0.0397 |
| **Department** |  |  |  |  |
| Allergy/Immunology | 12811 (11.00%) | 9643 (10.35%) | <0.001 | 0.021 |
| Cardiology | 18532 (15.92%) | 10872 (11.66%) | <0.001 | **0.1162\*** |
| Craniofacial Surgery | 694 (0.60%) | 564 (0.61%) | 0.7879 | 0.0012 |
| Cardiothoracic Surgery | 1881 (1.62%) | 1239 (1.33%) | <0.001 | 0.0227 |
| Dermatology | 1550 (1.33%) | 1187 (1.27%) | 0.2486 | 0.005 |
| Endocrinology | 12739 (10.94%) | 9290 (9.97%) | <0.001 | 0.0312 |
| Gastroenterology | 10864 (9.33%) | 11661 (12.51%) | <0.001 | **0.1094\*** |
| Hematology/Oncology | 7350 (6.31%) | 3342 (3.59%) | <0.001 | **0.1121\*** |
| Infectious Diseases | 1125 (0.97%) | 920 (0.99%) | 0.6276 | 0.0021 |
| Neurology | 11847 (10.17%) | 13610 (14.60%) | <0.001 | **0.1465\*** |
| Neonatology | 1329 (1.14%) | 1366 (1.47%) | <0.001 | 0.0305 |
| Neurosurgery | 4119 (3.54%) | 3665 (3.93%) | <0.001 | 0.0214 |
| Nephrology | 6152 (5.28%) | 6946 (7.45%) | <0.001 | 0.097 |
| Orthopedics | 13942 (11.97%) | 8209 (8.81%) | <0.001 | 0.0975 |
| Plastic Surgery | 304 (0.26%) | 225 (0.24%) | 0.3725 | 0.0039 |
| Pulmonology | 1812 (1.56%) | 2425 (2.60%) | <0.001 | 0.0845 |
| Sports Medicine | 298 (0.26%) | 153 (0.16%) | <0.001 | 0.0182 |
| Surgery (General) | 3071 (2.64%) | 1823 (1.96%) | <0.001 | 0.0425 |
| Urology | 17474 (15.01%) | 19321 (20.73%) | <0.001 | **0.1603\*** |
| Other Departments | 395 (0.34%) | 351 (0.38%) | 0.1533 | 0.0064 |
| **Days Until Scheduled Appointment** | 48.06 [47.68-48.44] | 82.28 [81.74-82.82] | <0.001 | **0.5180\*** |
| **Appointment Length** | 25.05 [24.98-25.11] | 26.45 [26.38-26.52] | <0.001 | **0.1251\*** |
| **Referral Required** | 16837 (14.46%) | 14858 (15.94%) | <0.001 | 0.0421 |
| **Overbooked Timeslot** | 12302 (10.56%) | 5602 (6.01%) | <0.001 | **0.1482\*** |
| **Same Day Appointment** | 9926 (8.52%) | 829 (0.89%) | <0.001 | **0.2734\*** |
| **Canceled Appointment Ratio** | 20.40% [20.29%-20.51%] | 29.07% [28.93%-29.22%] | <0.001 | **0.4544\*** |
| **No-Show Appointment Ratio** | 3.65% [3.62%-3.69%] | 5.12% [5.07%-5.17%] | <0.001 | **0.2249\*** |
| **Guardian Gender (% Male)** | 14325 (22.98%) | 7380 (19.86%) | <0.001 | 0.0742 |
| **Employment Status** |  |  |  |  |
| Full-Time | 58784 (50.48%) | 31562 (33.86%) | <0.001 | **0.3324\*** |
| Part-Time | 7903 (6.79%) | 4758 (5.11%) | <0.001 | 0.0669 |
| Student | 1814 (1.56%) | 977 (1.05%) | <0.001 | 0.0412 |
| Unemployed | 32678 (28.06%) | 23121 (24.81%) | <0.001 | 0.0725 |
| **Insurance (% Medicaid)** | 50497 (43.37%) | 35233 (37.80%) | <0.001 | **0.1123\*** |
| **Copay Due at Visit** | 8.07 [7.98-8.16] | 6.29 [6.2-6.38] | <0.001 | **0.1146\*** |
| **Maximum Temperature** | 65.23 [65.12-65.33] | 63.94 [63.82-64.06] | <0.001 | 0.0699 |
| **Minimum Temperature** | 47.41 [47.31-47.51] | 46.35 [46.24-46.47] | <0.001 | 0.0615 |
| **Temperature** | 56.19 [56.09-56.29] | 55.03 [54.92-55.15] | <0.001 | 0.0673 |
| Wind Chill | 27.32 [27.21-27.43] | 25.96 [25.83-26.09] | <0.001 | **0.1008\*** |
| Precipitation | 0.13 [0.13-0.13] | 0.13 [0.13-0.13] | 0.0235 | 0.01 |
| Wind Speed | 10.38 [10.36-10.4] | 10.42 [10.4-10.44] | 0.0057 | 0.0121 |
| Cloud Cover | 52.53 [52.35-52.72] | 53.79 [53.59-54] | <0.001 | 0.0395 |
| Relative Humidity | 67.26 [67.19-67.34] | 67.45 [67.37-67.53] | 0.0011 | 0.0142 |
| Weather - Clear | 29065 (24.96%) | 22124 (23.74%) | <0.001 | 0.0283 |
| Weather - Rain | 56191 (48.26%) | 45990 (49.34%) | <0.001 | 0.0218 |
| Weather - Overcast | 35816 (30.76%) | 30136 (32.33%) | <0.001 | 0.0341 |
| Weather - Partially cloudy | 51561 (44.28%) | 40942 (43.93%) | 0.1066 | 0.0071 |

**Table 2.** Model performance metrics on the 2019 validation data.

|  | Precision | Recall | F1-score |
| --- | --- | --- | --- |
| **All Features** |  |  |  |
| Negative Class | 0.83 | 0.75 | 0.79 |
| Positive Class | 0.82 | 0.88 | 0.85 |
| Accuracy |  |  | 0.82 |
| **Top 5 Features** |  |  |  |
| Negative Class | 0.78 | 0.74 | 0.76 |
| Positive Class | 0.81 | 0.84 | 0.82 |
| Accuracy |  |  | 0.80 |

**Table 3.** Model training on the entire dataset followed by application and evaluation on individual racial and ethnic groups.

| Racial/Ethnic Group | 2015-2018 (# of Patients and Total Percent) | 2019 (# of Patients and ROC AUC) | 2020 (# of Patients and ROC AUC) |
| --- | --- | --- | --- |
| Caucasian | 177, 375 (93%) | 67,559 (AUC: 0.89) | 47,495 (AUC: 0.89) |
| Black | 5,644 (3.0%) | 1,999 (AUC: 0.84) | 1,466 (AUC: 0.83) |
| Two or More Races | 4,570 (2.4%) | 1,933 (AUC: 0.75) | 1,408 (AUC: 0.83) |
| Hispanic/Latino | 2,386 (1.2%) | 1,095 (AUC: 0.79) | 731 (AUC: 0.81) |
| Asian American | 1,073 (0.6%) | 568 (AUC: 0.83) | 372 (AUC: 0.85) |
| Native American | 204 (0.1%) | 69 (AUC: 0.84) | 84 (AUC: 0.78) |

### Evaluation of Model Generalizability and Adaptability to unknown Healthcare Needs

We also wanted to understand if our model was adaptable to the changing dynamics of healthcare needs and initiatives, such as those driven by the Coronavirus Disease 2019 (COVID-19), which created an increase of no-contact telehealth appointments. The evaluation of our holdout dataset (2020), which contained 35% telehealth visits, revealed that our model provided superior predictions across all features (AUC: 0.90) and the topmost predictive features (AUC: 0.88) (Figure 3). Again, we showed that the traditional “no-show rate” computed in the EMR system was significantly inferior to our integrative approach (AUC: 0.62). We further evaluated the top 5 features in the model, which revealed the same features as seen in the validation dataset (i.e., Demographics-8, Time-3, Hospital-5, Appointment-11, and Appointment-10) (Data Not Shown). Model robustness on the 2020 pediatric telemedicine dataset and the shared top features between the validation and external holdout dataset highlight the persistence of our machine learning model to generate accurate predictions of patient no-shows. Additionally, the preliminary data is from pediatric appointments, highlighting our algorithm’s ability to predict the no-show rate of the patient based primarily on external factors, such as transportation by the guardian/caregiver.

**Figure 3.**
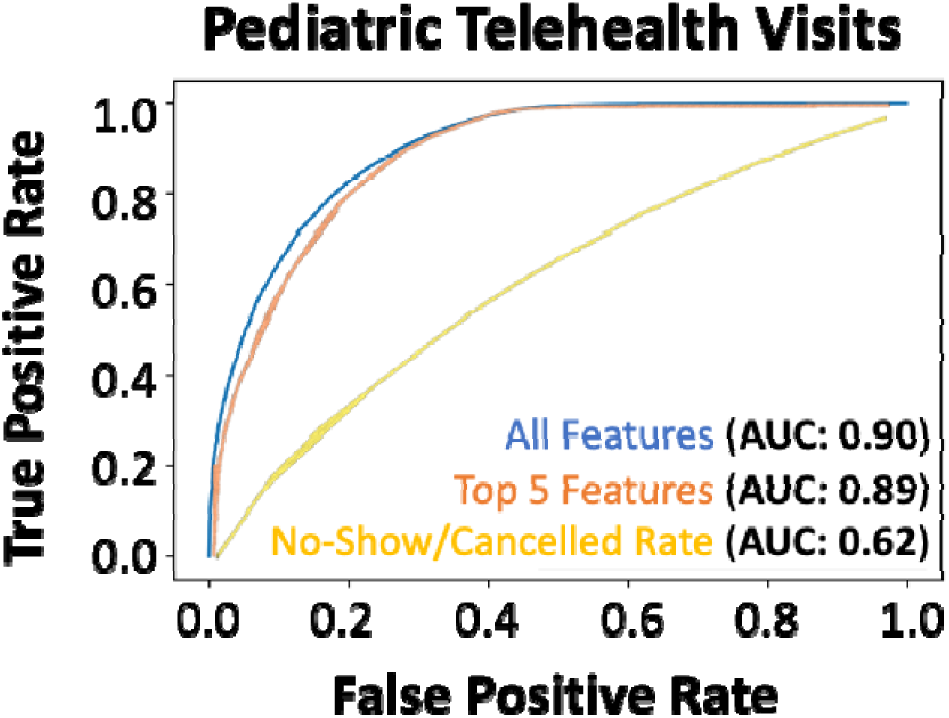
Performance of the machine learning (XGBoost) model for predicting no-shows in the external validation dataset. AUROC of xgboost machine learning model all features (blue line), only the top-5 features (orange line) and the direct no-show/cancel rate (yellow line) in predicting no-shows of the 2020 telemedicine dataset during the pandemic season.

For our system at WVU, we learned that strategic double booking might be a solution. Most of our clinics had 20 minutes visit slots, so for a 4 hours clinic session, our proposed double booking was focused towards the middle of the session with a limit of 2 per 4 hours and 3 for 8 hours. Some of our specialists had 30 minutes for a return visit and 1 hour for a new patient visit. For those schedules our strategy was to overbook a follow up visit in the new patient visit slot with a new patient who had waited over 3 months for the appointment as the data suggested decreased probability of patient adherence to a scheduled appointment after 3 months wait time. We strongly recommended not overbooking at the beginning or end of a clinic session to maintain the flow of the clinic as well as considering the providers efficiency during the clinic session. As the technology improves all these measures might even become second nature in clinic patient scheduling.

## Discussion

Our technology uses an AI-based machine-learning algorithm to predict the probability of individual patients not showing to an appointment on a given date and time. The algorithm uses a patient’s historical demographic data to proactively predict no-shows and employ strategic double booking to avoid disruptions and minimize costs to the clinic. Unlike other scheduling systems that employ advanced statistical techniques (such as logistic regression and Bayesian prediction), our platform leverages cutting-edge machine learning technology -- a scalable, distributed gradient-boosted decision tree algorithm utilizing minimal feature input for easy implementation across all healthcare systems -- that continues to learn and improve, thus increasing prediction accuracy over time.

Of the work that has been done to improve patient no-show prediction, many studies have failed to perform superiorly to the basic statistic of no-show rate displayed per patient [17]. Of the studies that performed better than the no-show rate basic statistic, most algorithms have not shown superior performance in predicting patient no-shows (i.e., AUC <0.85) and have also only been applied to very specific populations (i.e., a single population or visit type) [17]. While the current algorithms predicting patient no-shows reveal promise for clinical application [27, 28], there is currently no validated machine learning algorithm incorporated into an EMR to provide real-time predictions. Additionally, due to the limited scope of most no-show prediction algorithms, racial bias within the algorithm can result in up to a 30% increase in wait time for black patients [29]; optimizing care for patients, regardless of gender, race/ethnicity, and socioeconomic status, requires a demonstration of the algorithms stability across these conditions to promote healthcare equity.

Clinics currently utilize scheduling protocols that dictate how patients get scheduled for each clinic and within different specialties. There’s considerable variability per provider, per clinic, and sometimes even per site within a healthcare system. Epic Systems Corporation (Epic) Electronic Medical Record (EMR) has a basic statistic that displays the no-show rate per patient in the scheduling software, but there is little knowledge about the prospective performance of this information. However, this information is not currently utilized in improving the scheduling of appointments.

While the scheduling staff can see the historic no-show rate for each patient when they call for an appointment, they have no autonomy to actively modify scheduling to reduce patient no-shows and follow the guidance of clinic protocols. As such, commonly implemented approaches to avoid no-shows include appointment reminders and no-show fines, while approaches to reduce the impact of no-shows on the providers and the healthcare system include double booking. Double booking could offer an advantage to both the patient and provider if implemented in a strategic manner, including through combination with predictive AI algorithms. However, such automated “strategic double booking” would require a machine-learning algorithm capable of dynamically updating based on each patient’s likelihood of missing an appointment. For example, at WVU, strategic double booking was observed to be an effective approach. In clinics with 20-minute visit slots, double booking was typically concentrated in the middle of 4-hour sessions, with a common limit of two patients for a 4-hour session and three for an 8-hour session. For specialists with 30-minute return visits and 1-hour new patient appointments, follow-up visit slots were often overbooked with new patients who had been waiting over 3 months, as data suggested a decrease in adherence following such delays. Additionally, overbooking at the start or end of clinic sessions was generally avoided to maintain clinic flow and optimize provider efficiency. As the technology improves all these measures might even become second nature in clinic patient scheduling. Thus, there is an unmet need for generalizable machine-learning approaches to both predict and proactively address patient no-shows and their impact on the healthcare system.

## Limitations

Our study looked at the clinic population in the Appalachian region of west Virginia. Even though there was financial diversity within the state was taken into consideration, there might be factors that were not obvious in our results due to limited ethnic and racial diversity in the state of West Virginia. While our study provides valuable insights, it is important to acknowledge limitations such as potential data biases and the retrospective nature of the analysis. Importantly, the data used in this study are provided by a specific hospital or hospital system, and in pediatric population thus the generalizability of the research results may be limited. Future research can extend the data source to other hospitals and to other population and age ranges to cross-validate our results or could focus on prospectively collecting data and implementing interventions to evaluate their effectiveness in reducing patient no-show rates.

Our proposed solution of strategic overbooking, itself, has its limitation. It depends on the protocols shared with the call center to offer certain slots for overbooking. We believe that a model actively analyzing the patient no-show data and proposing slots in real time using the patient no-show history might be a better solution to take away any end user bias or human errors in interpreting the protocol implementation.

## Conclusions

No-shows in healthcare settings pose significant challenges for both providers and patients. Understanding the underlying factors driving these no-shows is crucial for developing effective interventions to reduce their occurrence and enhance clinic efficiency. This study proposes the implementation of a machine learning framework to predict patient no-shows, offering hospitals a proactive approach to optimize their outpatient appointment systems. By accurately anticipating potential no-show behavior, healthcare facilities can implement targeted strategies such as personalized appointment reminders, flexible scheduling options, and provider-specific interventions to mitigate the impact of no-shows on healthcare delivery. These proactive measures not only improve clinic efficiency but also enhance patient satisfaction, provider productivity and overall healthcare outcomes.

## Conflict of Interest

The authors declare that the research was conducted in the absence of any commercial or financial relationships that could be construed as a potential conflict of interest.

## Author Contributions

JN, TC and NY contributed to conception and design of the study. NY organized the database. QH and NY performed the statistical analysis and developed models. QH wrote the first draft of the manuscript. JN, TC, and NY wrote sections of the manuscript. All authors contributed to manuscript revision, read, and approved the submitted version.

## Funding

N/A

## Data Availability

All data produced in the present study are available upon reasonable request to the authors.

## Acknowledgments

The authors express their gratitude to all the patients, providers, administrators, clinic staff, schedulers and researchers at West Virginia University Hospitals who contributed and/or supported our research.

## Notes

### Competing Interest Statement

The authors have declared no competing interest.

### Funding Statement

This study did not receive any funding.

### Author Declarations

Institutional Review Board (IRB) of West Virginia University gave ethical approval for this work.

## References

Ahmadi, E., Garcia-Arce, A., Masel, D.T., Reich, E., Puckey, J., and Maff, R. (2019). A metaheuristic-based stacking model for predicting the risk of patient no-show and late cancellation for neurology appointments. IISE Transactions on Healthcare Systems Engineering 9, 272–291.

Alshammari, A., Almalki, R., and Alshammari, R. (2021). Developing a Predictive Model of Predicting Appointment No-Show by Using Machine Learning Algorithms. Journal of Advances in Information Technology 12, 234–239.

Alshammari, R., Daghistani, T., and Alshammari, A. (2020). The Prediction of Outpatient No-Show Visits by using Deep Neural Network from Large Data. (IJACSA) International Journal of Advanced Computer Science and Applications 11, 533–539.

Ansell, D., Crispo, J.a.G., Simard, B., and Bjerre, L.M. (2017). Interventions to reduce wait times for primary care appointments: a systematic review. BMC Health Serv Res 17, 295.

Chen, J., Goldstein, I.H., Lin, W.C., Chiang, M.F., and Hribar, M.R. (2020). Application of Machine Learning to Predict Patient No-Shows in an Academic Pediatric Ophthalmology Clinic. AMIA Annu Symp Proc 2020, 293–302.

Chong, L.R., Tsai, K.T., Lee, L.L., Foo, S.G., and Chang, P.C. (2020). Artificial Intelligence Predictive Analytics in the Management of Outpatient MRI Appointment No-Shows. AJR Am J Roentgenol 215, 1155–1162.

Daghistani, T., Alghamdi, H., Alshammari, R., and Alhazme, R.H. (2020). Predictors of outpatients’ no-show: big data analytics using apache spark. Journal of Big Data 7.

Ellis, D.A., Mcqueenie, R., Mcconnachie, A., Wilson, P., and Williamson, A.E. (2017). Demographic and practice factors predicting repeated non-attendance in primary care: a national retrospective cohort analysis. Lancet Public Health 2, e551–e559.

Fan, G., Deng, Z., Ye, Q., and Wang, B. (2021). Machine learning-based prediction models for patients no-show in online outpatient appointments. Data Science and Management 2, 45–52.

Festinger, D.S., Lamb, R.J., Marlowe, D.B., and Kirby, K.C. (2002). From telephone to office: intake attendance as a function of appointment delay. Addict Behav 27, 131–137.

Gier, J. (2017). Missed appointments cost the U.S. healthcare system $150B each year [Online]. Online: Healthcare Innovation. Available: https://www.hcinnovationgroup.com/clinical-it/article/13008175/missed-appointments-cost-the-us-healthcare-system-150b-each-year [Accessed 2023].

Hayhurst, C. (2019). No-show effect: Even one missed appointment risks retention [Online]. Online: Athenahealth. Available: https://www.athenahealth.com/knowledge-hub/financial-performance/no-show-effect-even-one-missed-appointment-risks-retention [Accessed 2023].

Liu, D., Shin, W.Y., Sprecher, E., Conroy, K., Santiago, O., Wachtel, G., and Santillana, M. (2022). Machine learning approaches to predicting no-shows in pediatric medical appointment. NPJ Digit Med 5, 50.

Marbouh, D., Khaleel, I., Al Shanqiti, K., Al Tamimi, M., Simsekler, M.C.E., Ellahham, S., Alibazoglu, D., and Alibazoglu, H. (2020). Evaluating the Impact of Patient No-Shows on Service Quality. Risk Manag Healthc Policy 13, 509–517.

Mohammadi, I., Wu, H., Turkcan, A., Toscos, T., and Doebbeling, B.N. (2018). Data Analytics and Modeling for Appointment No-show in Community Health Centers. J Prim Care Community Health 9, 2150132718811692.

Moharram, A., Altamimi, S., and Alshammari, R. (2021). "Data Analytics and Predictive Modeling for Appointments No-show at a Tertiary Care Hospital", in: 2021 1st International Conference on Artificial Intelligence and Data Analytics (CAIDA). (Riyadh, Saudi Arabia).

Qureshi, Z., Maqbool, A., Mirza, A., Iqbal, M.Z., Afzal, F., Kanubala, D.D., Rana, T., Umair, M.Y., Wakeel, A., and Shah, S.K. (2021). Efficient Prediction of Missed Clinical Appointment Using Machine Learning. Comput Math Methods Med 2021, 2376391.

Salazar, L.H.A., Leithardt, V.R.Q., Parreira, W.D., Fernandes, A.M.R., Barbosa, J.L.V., and Correia, S.D. (2021). Application of Machine Learning Techniques to Predict a Patient’s No-Show in the Healthcare Sector. Future Internet 14, 3.

Salazar, L.H.A., Parreira, W.D., Fernandes, A.M.R., and Leithardt, V.R.Q. (2022). No-Show in Medical Appointments with Machine Learning Techniques: A Systematic Literature Review. Information 13, 507.

Shameer, K., Johnson, K.W., Yahi, A., Miotto, R., Li, L.I., Ricks, D., Jebakaran, J., Kovatch, P., Sengupta, P.P., Gelijns, S., Moskovitz, A., Darrow, B., David, D.L., Kasarskis, A., Tatonetti, N.P., Pinney, S., and Dudley, J.T. (2017). Predictive Modeling of Hospital Readmission Rates Using Electronic Medical Record-Wide Machine Learning: A Case-Study Using Mount Sinai Heart Failure Cohort. Pac Symp Biocomput 22, 276–287.

Srinivas, S., and Salah, H. (2021). Consultation length and no-show prediction for improving appointment scheduling efficiency at a cardiology clinic: A data analytics approach. International Journal of Medical Informatics 145.

